# From Answers to Insights: Unveiling the Strengths and Limitations of ChatGPT and Biomedical Knowledge Graphs

**DOI:** 10.1101/2023.06.09.23291208

**Authors:** Yu Hou, Jeremy Yeung, Hua Xu, Chang Su, Fei Wang, Rui Zhang

## Abstract

Large Language Models (LLMs) have demonstrated exceptional performance in various natural language processing tasks, utilizing their language generation capabilities and knowledge acquisition potential from unstructured text. However, when applied to the biomedical domain, LLMs encounter limitations, resulting in erroneous and inconsistent answers. Knowledge Graphs (KGs) have emerged as valuable resources for structured information representation and organization. Specifically, Biomedical Knowledge Graphs (BKGs) have attracted significant interest in managing large-scale and heterogeneous biomedical knowledge. This study evaluates the capabilities of ChatGPT and existing BKGs in question answering, knowledge discovery, and reasoning. Results indicate that while ChatGPT with GPT-4.0 surpasses both GPT-3.5 and BKGs in providing existing information, BKGs demonstrate superior information reliability. Additionally, ChatGPT exhibits limitations in performing novel discoveries and reasoning, particularly in establishing structured links between entities compared to BKGs. To overcome these limitations, future research should focus on integrating LLMs and BKGs to leverage their respective strengths. Such an integrated approach would optimize task performance and mitigate potential risks, thereby advancing knowledge in the biomedical field and contributing to overall well-being.

## Introduction

In recent years, Large Language Models (LLMs) have exhibited exceptional performance across a diverse range of natural language processing tasks^1–3^. LLMs, such as GPT-3.5 and GPT-4, are powerful models trained on vast amounts of textual data, enabling them to generate human-like text and perform various language-related tasks^4^. These models have found applications in diverse domains, including chatbots, question-answering systems, and language translation, among others. Their ability to understand and generate text has sparked interest in exploring their potential to replace traditional knowledge resources.

Knowledge Graphs (KGs) serve as valuable repositories of structured information, and have gained significant attention due to their ability to represent and organize knowledge in a structured manner. They facilitate knowledge discovery, entity linking, and semantic querying, making them essential for various applications, including information retrieval, recommendation systems, and semantic search. In recent years, the field of biomedicine has witnessed the emergence of Biomedical Knowledge Graphs (BKGs) as a novel paradigm for managing large-scale and heterogeneous biomedical knowledge, which have garnered considerable interest in the biomedical community^5–10^. A BKG is a multi-relational graph or network that integrates, harmonizes, and stores biomedical knowledge acquired from single or multiple expert-derived knowledge sources. Over the past decade, substantial efforts have been dedicated to constructing BKGs by integrating diverse expert-curated knowledge bases^6,8,11–13^ and extracting knowledge from literature using natural language processing (NLP) techniques^14–16^. Consequently, numerous distinct BKGs have been developed^17–20^.

LLMs exhibit impressive language generation capabilities and have the potential to acquire knowledge from vast amounts of unstructured text. They can generate responses to questions and provide valuable insights. However, LLMs face several limitations when confronted with the biomedical domain, leading to issues like erroneous and inconsistent answers^21–23^. This study aims to evaluate the ChatGPT (a popular LLM) and BKG through comprehensive assessments encompassing querying existing biomedical knowledge, discovering novel knowledge, and providing reasoning capabilities. We shed light on the strengths and limitations of ChatGPT and existing KG, providing insights into their complementary roles in knowledge representation and utilization. Our findings contribute to the ongoing discussions surrounding the synergies and potential collaborations between LLMs and KGs in enhancing knowledge-driven applications.

## Methods

To evaluate the effectiveness of ChatGPT and Knowledge Graphs (KGs), we conducted a comprehensive assessment based on their performance in answering drug-related and dietary supplements (DS)-related questions, their knowledge discovery capabilities, and the comprehensiveness of the knowledge they provide. Specifically, we investigated ChatGPT’s ability to generate accurate and relevant responses to drug-related and DS-related queries and its potential for knowledge discovery by identifying hidden patterns and relationships.

### Performance Evaluation of ChatGPT and BKG in Question-Answering

The question-answering (Q&A) dataset was obtained by extracting questions (including their titles and contents) from the “Alternative Medicine” sub-category in Yahoo! Answers^24^. The questions were grouped into categories such as Adverse Effects, Background, Contraindication, Effectiveness, Indication, Interaction, Safety, Uncertain, Unclassified, and Usage. Initially, we randomly selected 5 questions from each group, resulting in a total of 50 questions. To collect responses from ChatGPT, we input the questions as prompts and record the generated answers. For retrieving answers from iDISK, an integrated knowledge graph focused on DS^25^, we followed a two-step process. Firstly, we identified the unique identifier of the subject and its corresponding relationship based on the question’s description. Next, we linked the identified object identifiers to their respective names and translated them into natural language. To evaluate the responses, we followed the LiveQA Track guidelines^26^ and assigned judgment scores on a scale of 1 to 4. Two experts who have medical backgrounds were introduced for manual scoring. A score of 1 indicated an incorrect response, 2 represented an incorrect but related answer, 3 denoted a correct but incomplete response, and 4 indicated a correct and complete answer. These scores were then transformed into a range of 0-3, with 0 indicating a poor or unreadable response, 1 representing a fair response, 2 signifying a good response, and 3 designating an excellent response. Using this scale, we calculated two metrics. Firstly, we computed the average score, which evaluated the first retrieved answer for each test question by converting the 1-4 level grades to the 0-3 scores^26,27^. Secondly, we measured the *succ*@*i* + metric, which represents the ratio of the number of questions with a score of *i* or higher (where *i* ranges from 2 to 4) to the total number of questions. For example, *succ*@2 + quantifies the percentage of questions that were answered by the conversational agent (CA) with at least a fair grade^26^. To assess the statistical differences in the performance of the three systems, first, the QQ-plot was performed to look at the normality of the data. Then a t-test was employed. The analysis is conducted using R 1.1 with the package “car^28^”.

### Performance Evaluation of ChatGPT and BKG in Knowledge Discovery

To test knowledge discovery capabilities between ChatGPT and existing KGs, we devised a prediction scenario that emulates the task of drug repurposing through link prediction within a knowledge graph. Our primary objective was to prompt ChatGPT to identify drugs or DSs that are not presently utilized for the treatment or prevention of Alzheimer’s Disease (AD) but possess the potential to be employed in such capacities. The specifically crafted prompts included:

1. Please provide the approved drugs that are not currently used to treat Alzheimer’s disease but are potentially available for the treatment of AD. And please give your rationale. (Drug)
2. Please provide which dietary supplements have the potential to treat/prevent Alzheimer’s disease. And please give your rationale. (DS)

Subsequently, we examined the answers generated by ChatGPT to determine if these answers met the following criteria: 1. whether they were already present in existing KGs (specifically, iBKH^29^ for drugs and ADInt^30^ for DSs); 2. whether they were documented in clinical trials; and 3. whether they were supported by existing literature.

### Performance Evaluation of ChatGPT and BKG in Knowledge Reasoning

To assess the comprehensiveness of ChatGPT’s knowledge base, we conducted an experiment to examine its capability in establishing associations between the proposed drug and DS candidates and AD. In our previous study, we investigated potential pharmaceuticals and DS for the treatment or prevention of AD using link prediction techniques^29,30^. Building upon these previous findings, our objective was to evaluate ChatGPT’s knowledge base by examining the associations it provides between these hypothetical drug/DS candidates and AD, as well as the corresponding references it offers to support these hypotheses. To accomplish this, we formulated scenario-based inquiries as follows:

1. Please show the association/linkage (direct link or indirect link) between Caryophyllus aromaticus and Alzheimer’s disease (AD) in a structured way (like a triplet). And please provide the reference for your finding.
2. Please show the association/linkage (direct link or indirect link) between Loperamide and Alzheimer’s disease (AD) in a structured way (like a triplet). And please provide the reference for your finding.

## Results

We conducted a comprehensive evaluation of the performance of three systems, namely GPT-4.0, iDISK, and GPT-3.5, in addressing a set of 43 questions. Table 1 demonstrated the performance of three systems in the Q&A task. To satisfy the assumptions of the t-test, we plotted the Q-Q Plot for the scores of each system and found all scores to be normally distributed. The comparison between ChatGPT (GPT-3.5) and KG revealed comparable performance (p-value: 0.20), indicating that both systems possess similar capabilities in answering questions. Notably, ChatGPT (GPT-4.0) demonstrated better performance than iDSIK and ChatGPT (GPT-3.5) (both p-value < 0.05) with an average score of 2.12, outperforming iDISK (average score: 1.64) and GPT-3.5 (average score: 1.44). In terms of providing references, iDISK surpassed ChatGPT (both GPT-4.0 and GPT-3.5) by offering the name of the database from which the data was retrieved. Conversely, ChatGPT (both GPT-4.0 and GPT-3.5) fell short in providing valid references, as the mentioned article names and/or authors were found to be fabricated.

**Table 1.** Performance of three systems in the Q&A task.

| Measurement | Systems |  |  |
| --- | --- | --- | --- |
|  | ChatGPT (GPT-4.0) | ChatGPT (GPT-3.5) | iDISK |
| Average Score | 2.12 | 1.44 | 1.64 |
| succ@1+ | 1.0 | 0.79 | 0.88 |
| succ@2+ | 0.70 | 0.51 | 0.61 |
| succ@3+ | 0.42 | 0.14 | 0.12 |
| For succ@*i*+ metric, a higher value indicates the better performance. |  |  |  |

Table 2 presents the results obtained from the conversation questions using ChatGPT (model GPT-3.5) to simulate a scenario related to drug repositioning. Upon conducting ten iterations of the question, we observed that ChatGPT (GPT-3.5) returned several identical responses. Specifically, Levetiracetam was suggested in 6 out of the ten iterations, while Lithium and Ibuprofen were each suggested in 5 iterations. Moreover, the majority of drugs suggested by ChatGPT (GPT-3.5) have been investigated for potential associations with Alzheimer’s Disease treatment in ClinicalTrials.gov. And, all of the drugs have been investigated in one or more scientific literature that explores their potential association with Alzheimer’s Disease. Upon examining these drugs within a comprehensive biomedical knowledge graph (iBKH), it was observed that nearly all the drugs suggested by ChatGPT (GPT-3.5) are present in iBKH with direct links to AD. For those drugs lacking direct links, the shortest distance between them and Alzheimer’s Disease is only 2, with reasonable paths existing, such as (Drug) - [Target_DG] - (Gene) - [Associate_DiG] - (AD). Analogous outcomes are observed in the employment of ChatGPT (GPT-3.5) to simulate exploratory scenarios involving novel DS that may potentially treat or prevent AD. In addition, all of the DSs recommended by ChatGPT (GPT-3.5) have been included in AD clinical trials. These DSs have been extensively researched for their correlation with AD, with findings documented in relevant academic publications. Furthermore, direct connections between the suggested DSs and AD can be identified within ADInt, a comprehensive knowledge graph encompassing AD-related concepts and various potential interventions. The recorded pathway in ADInt is denoted as (DS) - [Treats/Prevents] - (AD). Additionally, we executed a parallel experimental manipulation employing ChatGPT (GPT-4.0). Despite the enhanced diversity of the recommended drugs, they remained present in established clinical trials and literature. Concurrently, the associated pathway records persisted within the extant knowledge graph. (The detailed information was shown in the supplementary table 1.)

**Table 2.**
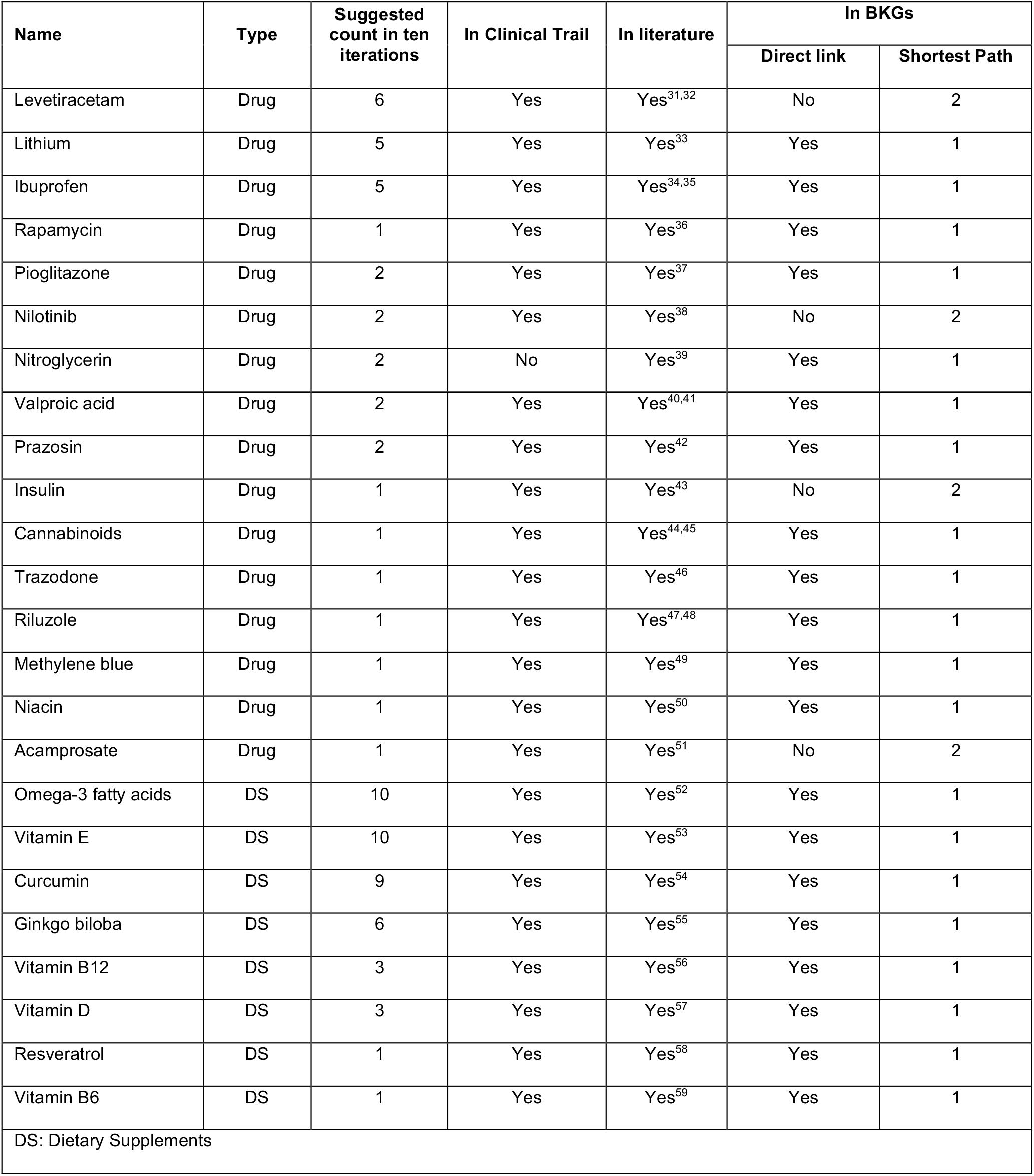

Figure 1 presents the responses of ChatGPT (GPT-3.5) and ChatGPT (GPT-4.0) to the devised scenario query 1, respectively. An examination of their replies reveals that ChatGPT endeavored to establish a structured connection between the hypothetical DS (Caryophyllus aromaticus) and Alzheimer’s disease (AD). Nonetheless, upon verification of the references provided by ChatGPT, it becomes evident that these citations are not authentic. Similar outcomes were observed during the investigation of novel drugs in the designed scenario 2. Figure 2 illustrates the attempts made by ChatGPT (GPT-3.5) and ChatGPT (GPT-4.0) to present structured links between the hypothetical drug (Loperamide) and AD. The connection offered by GPT-3.5 continues to be unsuccessful in validating the authenticity of the provided reference. Moreover, the response generated by GPT-4.0 indicates that it could not establish any potential direct or indirect association between Loperamide and AD.

**Figure 1.**
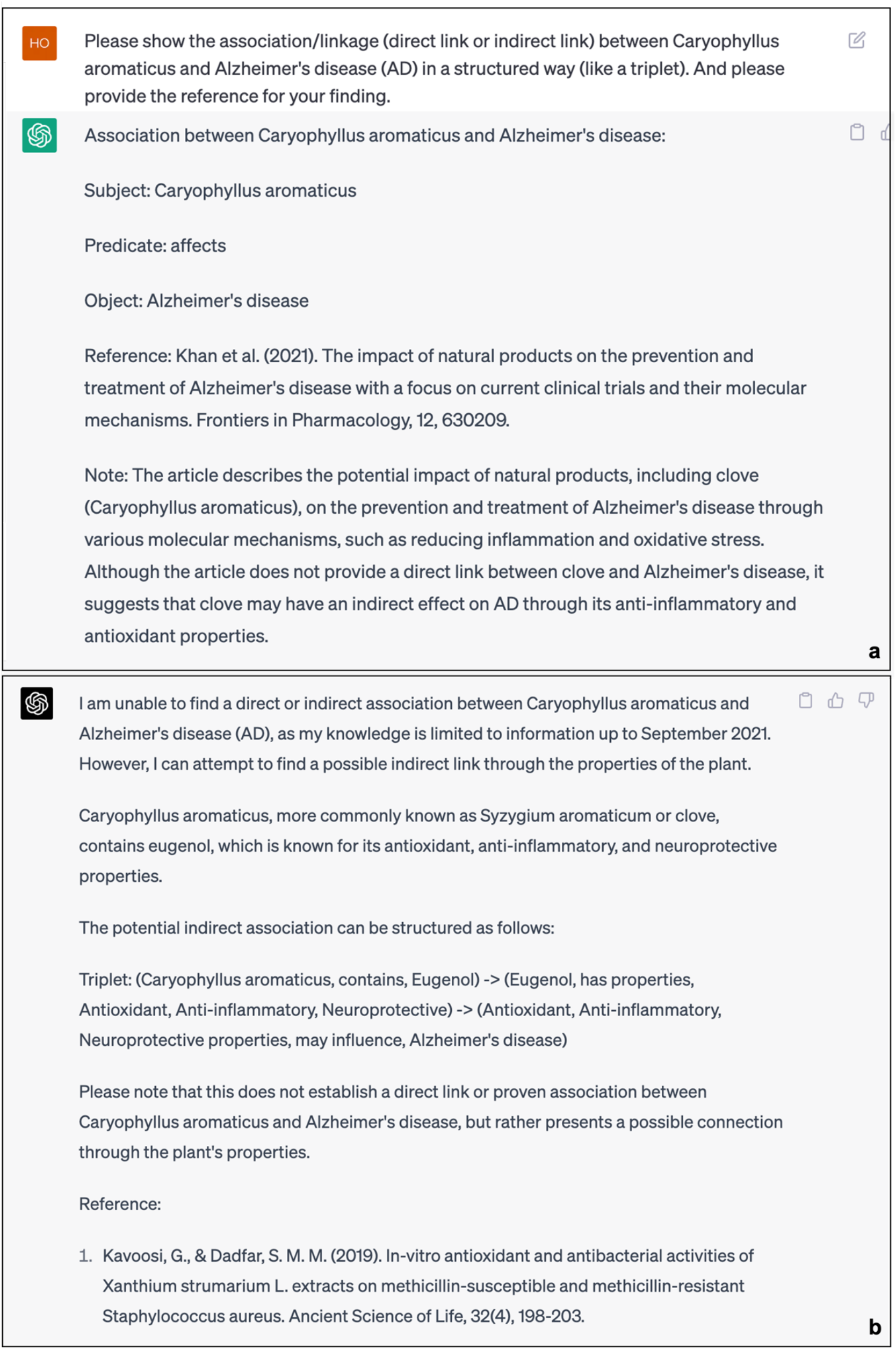
Responses of ChatGPT (GPT-3.5 for Fig. a and GPT-4.0 for Fig. b) to the Knowledge Reasoning task (DS)

**Figure 2.**
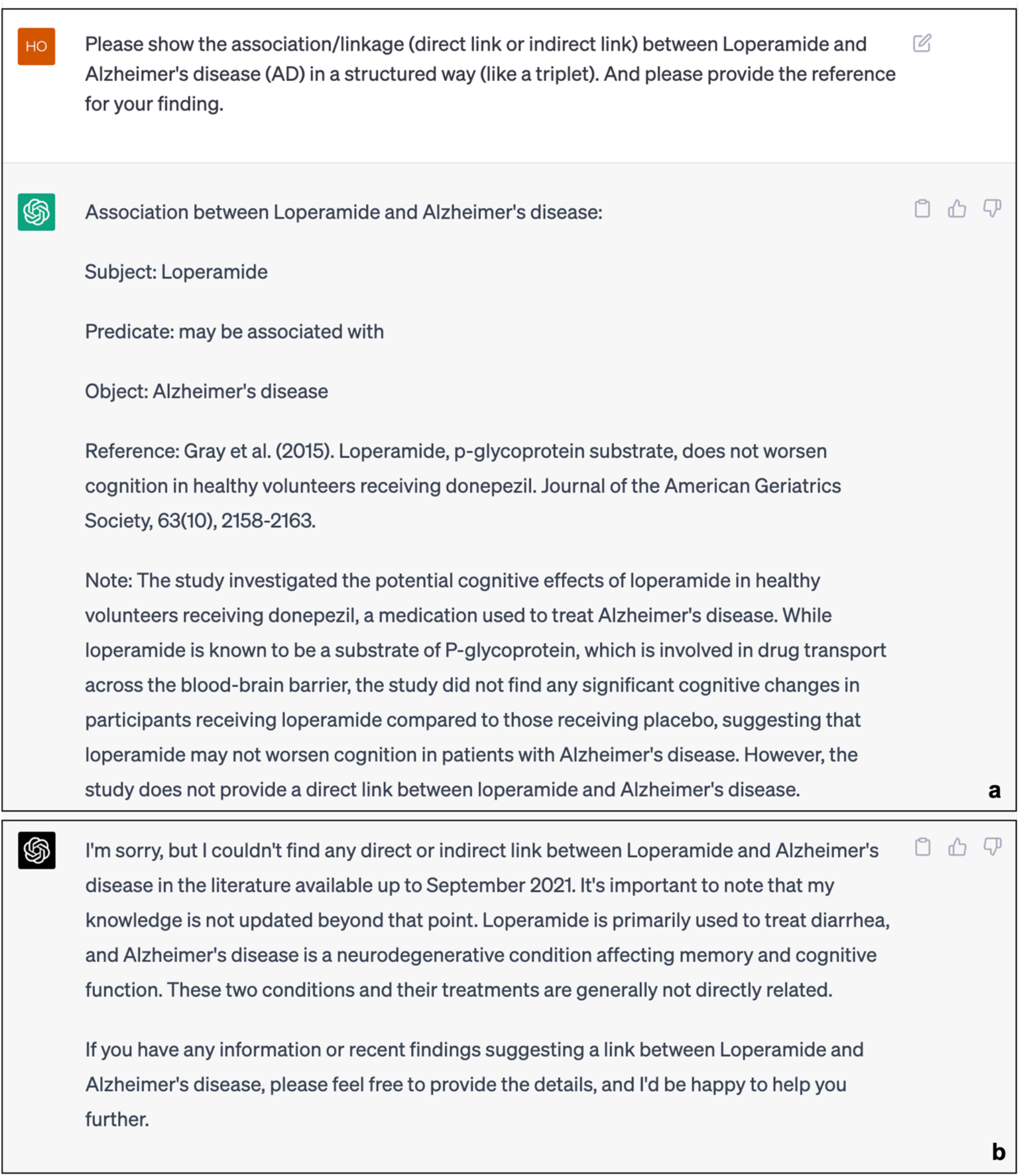
Responses of ChatGPT (GPT-3.5 for Fig. a and GPT-4.0 for Fig. b) to the Knowledge Reasoning task (Drug)

## Discussion

During the process of querying existing information, both ChatGPT (GPT-3.5) and BKG exhibited comparable capabilities, demonstrating their effectiveness in providing relevant information based on user queries. However, with the introduction of ChatGPT (GPT-4.0), a notable improvement in performance was observed compared to GPT-3 and BKG. The enhanced capabilities of ChatGPT (GPT-4.0) allowed for more accurate and comprehensive responses, surpassing the performance of both GPT-3 and KGs in this context. However, when it comes to the reliability of information sources, KG exhibited a clear advantage. KGs are built upon curated and structured knowledge from trusted sources, ensuring the reliability and accuracy of the information contained within them. This advantage stems from the rigorous data collection and validation processes employed in constructing BKG. On the other hand, ChatGPT’s responses are generated based on patterns and associations learned from a vast amount of text data, which may include both reliable and unreliable sources. As a result, while ChatGPT can provide quick responses, there may be a higher risk of encountering misinformation or inaccuracies compared to BKG. Therefore, when considering the reliability and trustworthiness of the information provided, KGs offer a more dependable and robust solution. Consequently, our findings highlight the potential benefits of integrating knowledge graph-based approaches with ChatGPT to enhance its domain-specific knowledge and overall performance in specialized applications. Further research is required to explore the feasibility of this integration and its implications on the efficacy of ChatGPT in diverse domains.

In addition, we discovered that ChatGPT is unable to perform the novel finding task based on existing knowledge, which is a critical limitation when considering its application in scientific research and discovery. In the second experiment, we attempted to simulate drug repurposing using ChatGPT as a means to generate innovative insights. The results of this experiment revealed that ChatGPT primarily provided outputs that were derived from pre-existing information. This information could either be directly queried within a knowledge graph or easily found in relevant resources, suggesting that ChatGPT’s capacity for generating truly novel findings is limited. These outcomes can be attributed to the underlying training data and architecture of ChatGPT, which is designed to draw upon its vast knowledge base to produce contextually relevant and coherent responses, rather than extrapolate new insights or hypothesize potential connections. This limitation highlights the need for developing advanced AI models that can not only process and comprehend existing knowledge but also deduce novel findings by identifying hidden patterns and relationships.

Our investigation revealed that ChatGPT exhibits limitations in its ability to establish a structured link between two entities based on existing knowledge. In our third experiment, we assessed the performance of ChatGPT in comparison to the BKG with respect to the establishment of relationships between entities. The results demonstrated that ChatGPT was unable to provide a structured link between two entities as effectively as the knowledge graph, underscoring its shortcomings in this specific task. Furthermore, the credibility of the results returned by ChatGPT emerged as a significant concern. Our findings indicated that the accuracy of its responses necessitates further verification, as the information provided by ChatGPT may not always be reliable. In the third experiment, it became evident that none of the references furnished by ChatGPT were genuine, casting doubt on the trustworthiness of the information it generated. These observations highlight the need for rigorous validation and verification mechanisms when employing ChatGPT for tasks that require high levels of accuracy and credibility. Consequently, future research should explore strategies to enhance the reliability of ChatGPT’s outputs, such as incorporating external validation sources or refining its training data to improve its capacity to provide accurate and credible information.

Our study assesses the capabilities of ChatGPT and existing BKGs in question answering, knowledge discovery, and knowledge reasoning. While ChatGPT with GPT-4.0 outperformed both GPT-3.5 and BKGs (both are comparable) to provide existing information; BKGs exhibited a clear advantage in terms of information reliability over both GPT models. Our findings revealed limitations in ChatGPT’s ability to perform novel discoveries based on existing knowledge. Furthermore, our investigation highlighted ChatGPT’s limitations in providing reasoning for knowledge discovery (e.g., establishing structured links between entities compared to BKGs). In conclusion, future investigations should prioritize the development of methodologies that integrate LLMs and BKGs, allowing researchers to harness the unique capabilities of each approach. This holistic approach would not only optimize task performance but also enable mitigating potential risks, thus advancing knowledge in the biomedical field and contributing to the overall well-being of individuals.

## Data Availability

All data produced are available online

## Funding

Research reported in this publication was supported by the National Institutes of Health (NIH)/National Institute On Aging (NIA) under Award Number R01AG078154 (PI: RZ) and the NIH/National Center For Complementary & Integrative Health (NCCIH) under Award Number R01AT00945 (PI: RZ).

## Author Contributions

YH and RZ conceived the study design and wrote the initial draft of the manuscript. YH implemented the experiments of the study. All authors contributed to the production of the final manuscript. JY contributed to the statistics. RZ, HX, CS and FW advised this project. All authors contributed to the production of the final manuscript.

## Notes

### Competing Interest Statement

The authors have declared no competing interest.

### Funding Statement

This study was funded by the National Institutes of Health (NIH)/National Institute On Aging (NIA) and NIH/National Center For Complementary & Integrative Health (NCCIH).

